# Quality of care for chronic conditions: Identifying specificities of quality aims based on scoping review and Delphi survey

**DOI:** 10.1101/2024.04.05.24305374

**Authors:** Grace Marie V. Ku, Willem van de Put, Deogratias Katsuva, Mohamad Ali Ag Ahmed, Megumi Rosenberg, Bruno Meessen

**Author notes:** Corresponding author: Grace Marie V. Ku Department of Public Health, Institute of Tropical Medicine Nationalestraat 155, 2000 Antwerp, Belgium.

## Abstract

**Background:** There is a need to implement good quality chronic care to address the ballooning burden of chronic conditions affecting all countries globally. However, to our knowledge, no systematic attempts have yet been made to define and specify aims for chronic care quality.

**Objective:** We conducted a scoping review and Delphi survey to establish and validate a comprehensive specification of chronic care quality aims.

**Methodology:** The Institute of Medicine’s (IOM) quality of care definition and aims was utilised as our base. We purposively selected scientific and grey literature that have acknowledged and unpacked the plurality of quality in chronic care and which proposed/made use of frameworks and studied their implementation or investigated minimum two IOM care quality aims and their implementation. We critically analysed the literature deductively and inductively. We validated our findings through Delphi survey involving international chronic care experts, mostly coming from/have expertise on low-and-middle-income countries.

**Results:** We considered the natural history of chronic conditions and the journey of a person with chronic condition to define and identify aims of chronic care quality. We noted that the six IOM aims apply but with additional meanings. We identified a seventh aim, continuity, which relates well to the issue of chronicity. Our panellists agreed with the specifications. Several provided contextualised interpretations and concrete examples.

**Conclusions:** Chronic conditions pose specific challenges underscoring the relevance of tailoring quality of care aims. Operationalization of this tailored definition and specified aims to improve, measure and assure quality of chronic care can be next steps.

## BACKGROUND

Chronic conditions, whether non-communicable or infectious, are broadly defined to last one year or more and require ongoing medical attention and/or limit activities of daily living [1]. These raise particular issues in terms of quality of care: persons with chronic conditions (PwCC) should be offered a seamless journey through time in the healthcare system, across services, providers, levels of care, etc [2–5]. More fundamentally, although healthcare providers (HCP) are expected to deliver healthcare, having a chronic condition means that PwCC are in-charge of their own health on a day-to-day basis. Daily decisions they make have a huge impact on their own health outcomes and quality of life [6]. Thus, chronic care should: include disease prevention and medication prescription activities; give focus on disability limitation, rehabilitation [7], and palliative care; and involve, enable and engage the PwCC (and their families) in taking care of the condition, controlling risks and promoting well-being (self-care) [8] considering not only biomedical but also psychosocial aspects *to adapt* and self-manage in the face of social, physical, and emotional challenges [9,10]. It is likewise crucial to acknowledge the reality of multimorbidity.

Although the above are delineated in standards of care for specific chronic conditions and are recognized in different models of chronic care, we did not find any established ready-to-use definition of what would be considered good quality chronic care [11]. Also, while there has been considerable work around a ‘global’ definition of quality of care and its core ‘aims’ over the last decades [12–15], there has been no systematic attempts made yet, to the best of our knowledge, to specify quality aims for chronic care. Having a tailored definition would empower the many and various actors – policy makers, health financiers/purchasing agencies, healthcare/service providers, healthcare regulators, accreditation agencies, researchers, trainers, PwCC themselves, etc – who are committed to achieve “good quality chronic care” and/or have the mandate to implement specific quality-enhancing interventions for chronic care. And while the need to deliver good quality chronic care to address the increasing burden of chronic conditions applies in all settings, it is highest in low-income settings.

This paper builds on a larger study commissioned by the World Health Organization (WHO). The request was to produce a comprehensive conceptualization of “quality health services for chronic conditions” that can be used by actors considering interventions to improve health services for chronic conditions, in this case, purchasing arrangements as an instrument for improvement, with a particular attention to policy needs of low- and middle-income countries (LMICs). As a first step, we created a framework for quality of chronic care. In this paper, we concentrate on the specification of chronic care quality aims, with the understanding that agreeing on the goals is a prerequisite before looking into determinants that can affect achieving these.

### Conceptual Issues

As a starting point, we adopted the definition of quality of care put forward by the Institute of Medicine (IOM)[13–15]: *Quality of care is the degree to which health services for individuals and populations increase the likelihood of desired health outcomes and are consistent with current professional knowledge*, noting that this generic definition needs to be contextualized and the outcomes to be determined.

While we are aware that different documents/reports have extended or reorganized these, we likewise took the IOM’s six aims as our base: effectiveness; efficiency; safety; equity; accessibility, timeliness, affordability; and person-centredness.

With the ideal that quality improvement should focus on the results that matter most to various actors (patients and their families, healthcare providers, regulators, decision makers, etc), we defined ‘aim’ as any broad category of importance with intrinsic value, as a desired final outcome that is achieved to denote that care is of good quality.

## METHODOLOGY

We reviewed relevant literature and convened international stakeholders of chronic care and quality in a Delphi survey.

We conducted scoping review following PRISMA extension guidelines [16] to systematically identify available information on quality of care for chronic conditions, identifying key concepts. We selected works that have acknowledged and unpacked the plurality of quality in chronic care, and which proposed/made use of frameworks or looked into two or more IOM aims of care quality, and studied or demonstrated implementation. The scoping review protocol is available from https://www.itg.be/en/research/research-themes/quality-of-care-for-chronic-conditions.

### Scientific publications

On February 2, 2022, search for scientific publications was conducted in the PubMed and Science Direct data bases using specific search terms: *‘chronic condition’/’chronic illness’/’chronic disease’; ‘quality of healthcare’; ‘innovative care for chronic conditions’; ‘chronic care model’; ‘quality criteria’; ‘quality indicators’*; specific chronic conditions considered among top drivers of chronic disease burden [17] (*‘ischaemic heart disease’, ‘hypertension’ and ‘stroke’; ‘diabetes mellitus’; ‘chronic kidney disease’; ‘lung cancer’; ‘HIV/AIDS’; ‘chronic obstructive pulmonary disease’ and ‘bronchial asthma’*) and additional conditions as suggested by the WHO team *(‘chronic musculoskeletal conditions’; ‘chronic skin disease’*); and criteria: *written in English or French; publication years 2002-2021; among humans*.

### Other literature and documents

Search for grey/other literature (policies, circulars, publications not available from scientific search engines) were conducted using similar keywords but including general quality of care documents, and with broader year limitations (1999-2022) in the Google search engine. Additionally, contacts from the WHO and healthcare regulatory agencies, organizations with chronic disease programs/projects, and various Ministries of Health and/or connected agencies were requested to share any documents they have produced as related to quality of care, specifically for chronic conditions.

### Literature sifting

Scientific publications were sifted through Rayyan (www.rayyan.ai). This was done systematically by minimum two members of the research team. A third researcher resolved any disagreements amongst the two, as needed. Scientific publications were initially screened through the titles. Abstracts of the chosen documents were individually reviewed. Full articles were scrutinized and selected; only documents that are relevant to this study were included in the final selection.

Grey literature and other documents were purposively collected.

### Data extraction and analysis

Data retrieval was systematically initiated by at least one of the members of the research team and verified by a different member. We critically analysed the literature. We made use of deductive approaches based on the IOM quality aims, and inductive approaches to identify any additional quality aims; utilising our definition of ‘aim’ to guide both. Narrative synthesis of retrieved information was done. We brought forward concepts related to aims of good quality chronic care. We note that quality aims were not always explicitly stated. We then deduced the aims based on our critical analysis of the text using the IOM aims as our base. We also identified any quality aim not included in the IOM proposition. Furthermore, analysis was reflective and iterative, going back to the literature as we identified additional concepts.

### Delphi survey

We invited 52 respondents representing various stakeholders of chronic care and quality (including PwCC and their carers) from all over the world. We conducted two rounds of the Delphi survey via an online application, Mesydel (https://mesydel.com/en). The first step was to arrive at an agreement over our scoping review findings and to propose financing mechanisms to improve quality of chronic care. The second round was to fine-tune purchasing arrangements. Our findings on financing for quality healthcare will be presented in a separate paper. For this paper, we concentrated on Round 1 results. We synthesised and critically analysed the responses, reflecting on our scoping review findings. The respondents provided rich justification why specific chronic care quality aims should be included.

### Ethical considerations

The Delphi survey protocol and an amendment thereto were reviewed and approved by the Institute of Tropical Medicine, Antwerp Institutional Review Board (protocol number 1627/22). Briefing sessions, scheduled in two moments to accommodate time differences, were conducted to orient prospective participants to the study (https://www.youtube.com/watch?v=A7FLpdIB0xs&t=1s). Full informed consent was obtained prior to participation.

## RESULTS

We retrieved 15,215 scientific articles and retained 48 [18–65](Figure 1). The study designs were: 17 reviews; 12 implementation research (quality improvement and/or model implementation); nine cross-sectional/surveys; three randomized controlled trials; three qualitative and mixed methods; two case studies; one qualitative study; and one position statement. Eighteen are specific for certain chronic conditions (diabetes=5, cardiovascular diseases including hypertension and stroke=5; HIV/AIDS=2; chronic obstructive pulmonary disease=2; chronic kidney disease=2; osteoarthritis=1; and cancer=1). Some targeted specific groups (elderly=5, children=1, female=1, informal caregiver=1). Forty-six propose and implement or demonstrate implementations of various models of quality of care, mostly in high income countries (n=31), five in LMICs; South Africa=3, Haiti=1, not specified=1), and the rest (n=10) said to be global/international. Majority (n=46) fit and consolidate the IOM definition of quality and two or more of the IOM care quality aims. A couple [46–47] consider Donabedian’s [66] elements of quality in healthcare.

**Figure 1.**
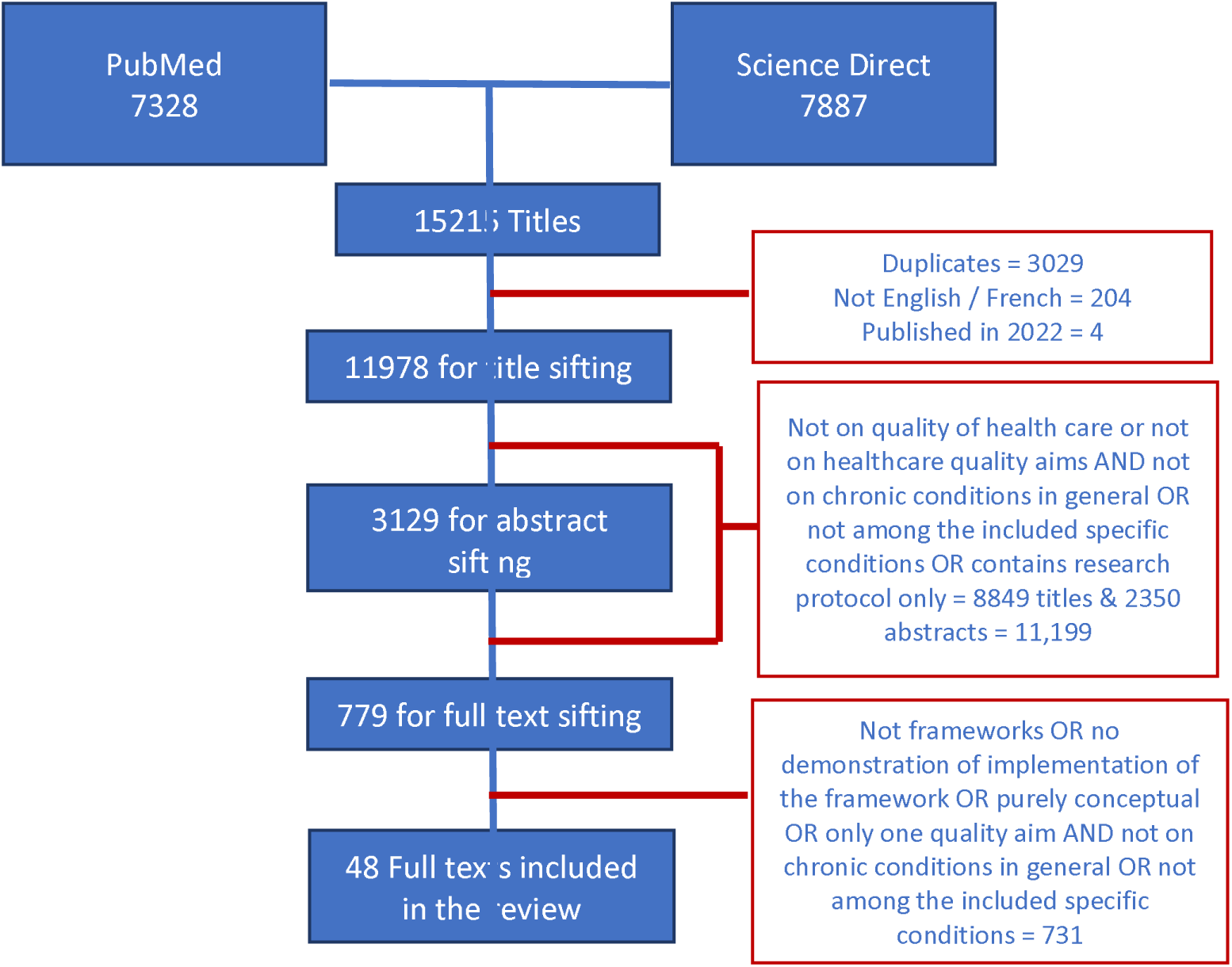
Sifting of retrieved scientific literature.

We retrieved 26 grey literature/documents from the WHO (n=10)[3,4,12,67-73); the EU Joint Action on Chronic Diseases and Healthy Ageing Across the Life Cycle (n=4)[74–77]; the IOM (n=3)[14–15,78]; the United States of America (USA) Agency for Health Care Research & Quality (n=2)[79–80]; and the rest coming from different agencies: two from the USA [81–82], and one document each from Australia [83], Belgium [84], Canada [85], Ireland [86], and the Philippines [87].

During data extraction, we noted that the scientific papers usually concentrate on a particular stage in what we deem to be the ‘journey’ of a person through healthcare, considering the natural history of (most) chronic conditions (e.g. risk-prevention/-control, follow-up, rehabilitation, etc). We thus went back to our selection to consciously extract additional information that would expound on the relevance of various stages in a continuum of care relevant to the identified aims. We will discuss the concept of the PwCC journey in a related paper.

Tables 1 and 2 provide overviews of the scientific articles and grey literature and chronic care quality aims identified.

**Table 1.**
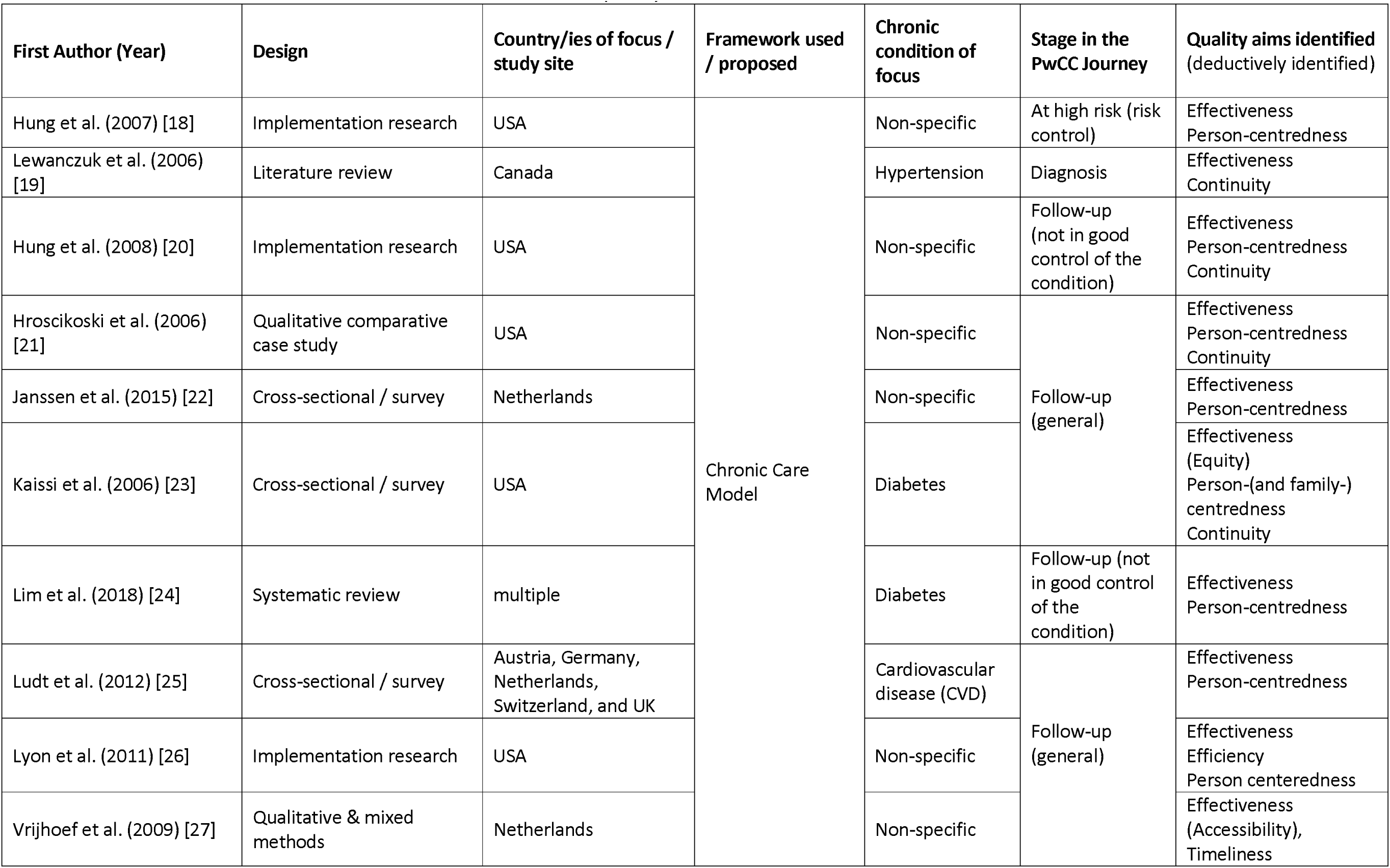
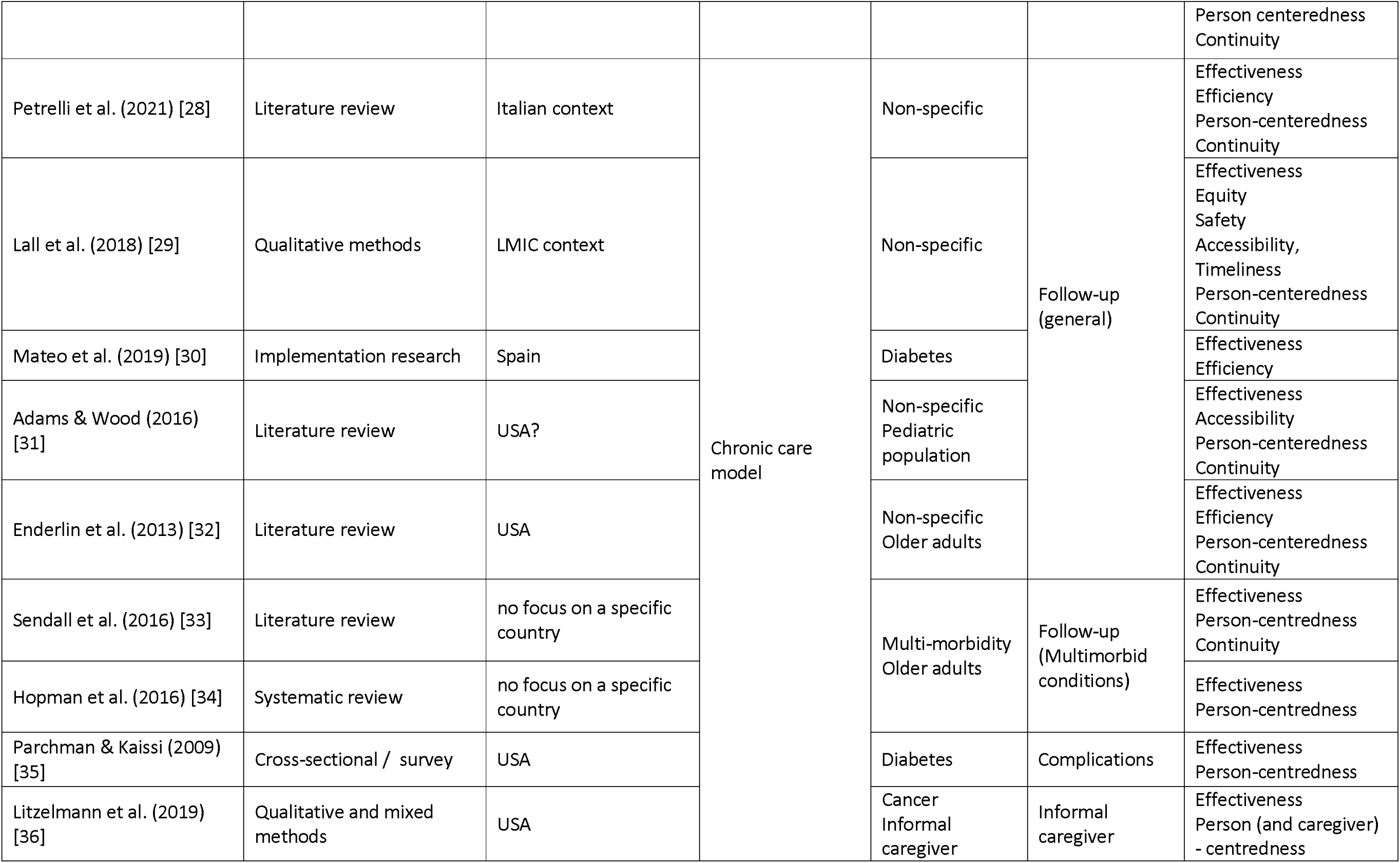
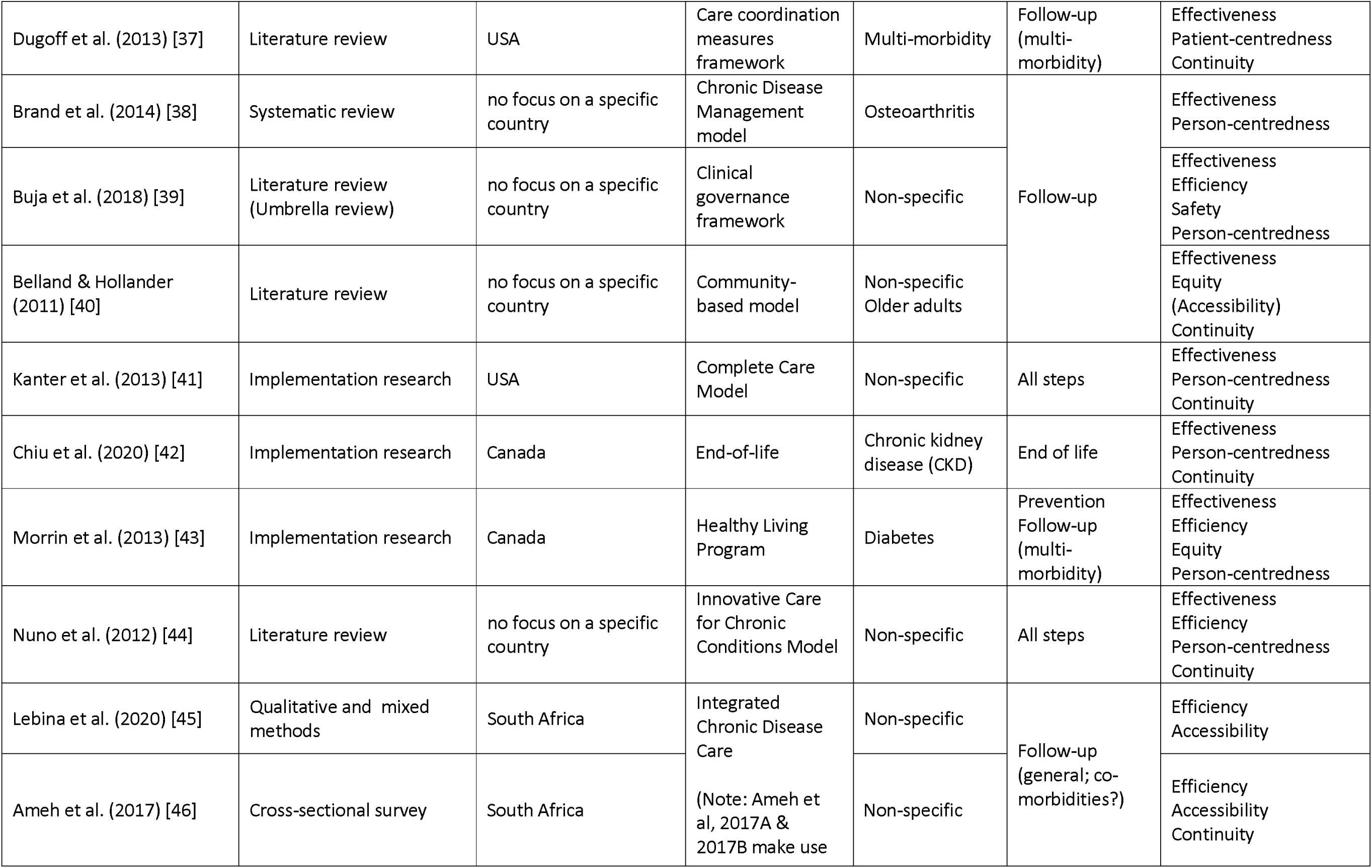
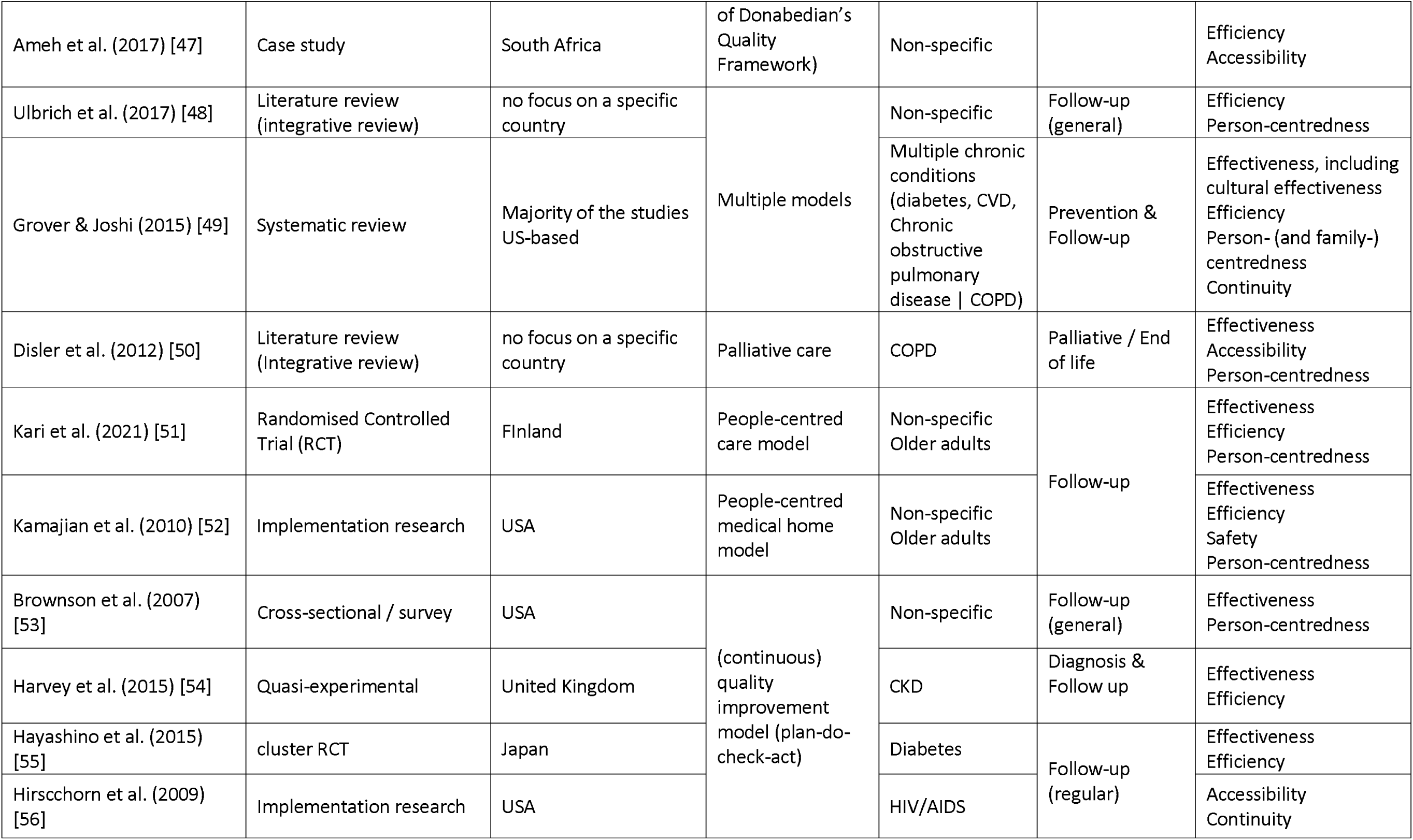
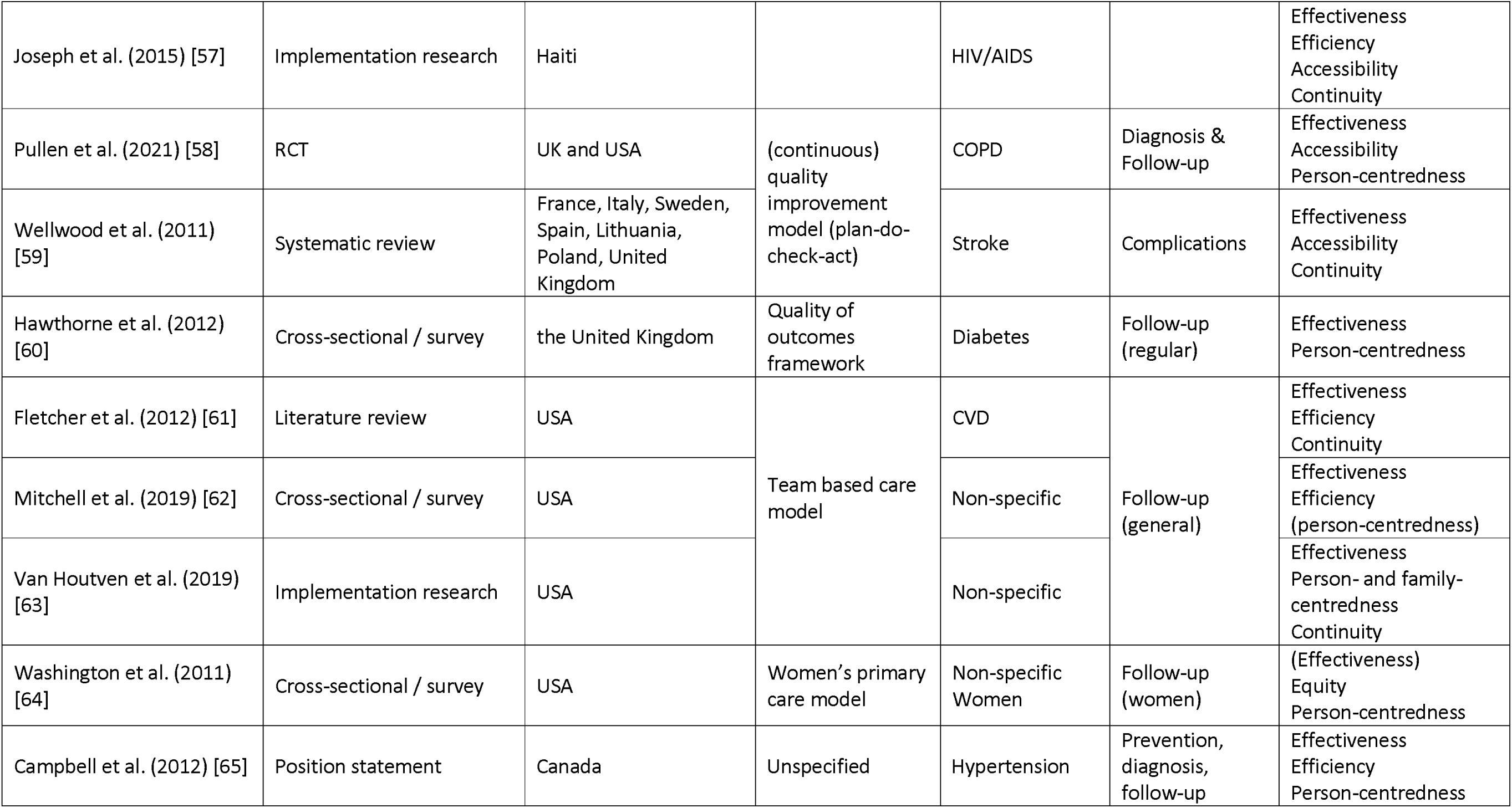
Overview of the scientific articles and chronic care quality aims identified.

**Table 2.** Overview of grey literature retrieved, setting, models/frameworks used, and quality aims identified.

| Author/editor/organization (year) | Setting | model(s) / framework(s) | Quality aims identified |
| --- | --- | --- | --- |
| World Health Organization (2013) [3] | Global | None | Effectiveness<br>Efficiency<br>Equity<br>Person-centredness<br>Continuity |
| World Health Organization (2016) [4] | Global | Integrated Care Models | Effectiveness<br>Efficiency<br>Person-centredness<br>Continuity |
| World Health Organization, Organisation for Economic Co-operation and Development, and the World Bank (2018) [12] | Global | Elements of health care quality | Effectiveness<br>Efficiency<br>Equity<br>Safety<br>Person-centredness<br>Timeliness<br>Continuity (as aim of integration) |
| Institute of Medicine (US) Committee on Quality of Health Care in America (2001) [14] | USA, but also used worldwide | IOM Quality aims | Safety<br>Effectiveness<br>Patient-centredness<br>Timeliness<br>Efficiency<br>Equity / equitability |
| Institute of Medicine (US) Committee on Quality of Health Care in America (2018) [15] | USA, but also used worldwide | IOM Quality aims | Effectiveness<br>Efficiency<br>Equity<br>Safety<br>Person-centredness<br>Accessibility, timeliness, and affordability (Continuity) |
| Escobar et al. (2013) [67] | Pan-American Region, but also used worldwide | None | Effectiveness |
| World Health Organization (2008) [68] | Global | None | Effectiveness<br>Efficiency<br>Person-centredness<br>Continuity |
| World Health Organization (2019) [69] | Global | None | Effectiveness<br>Efficiency |
| World Health Organization (2022) [70] | Global | Health systems performance | Effectiveness<br>Efficiency<br>Equity<br>Safety<br>Person-centredness (as aim of user experience)<br>Accessibility, timeliness, affordability |
| World Health Organization and the United Nations Children's Fund (2018) [71] | Global | Primary Health Care | Effectiveness<br>Person-centredness<br>Continuity |
| World Health Organization (2014) [72] | Global | NCD assessment guide | Effectiveness |
| World Health Organization (2002) [73] | Global | None | Effectiveness<br>Efficiency<br>Person-centredness<br>Continuity |
| Palmer et al. (2016) [74] | Europe | None | Effectiveness<br>Person-centredness<br>Continuity |
| EU Joint Action on Chronic Diseases and Healthy Ageing Across the Life Cycle (undated) [75] | Europe | practice quality evaluation tool | Effectiveness<br>Person-centredness |
| EU Joint Action on Chronic Diseases and Healthy Ageing Across the Life Cycle (undated) [76] | Europe | QCR tool | Effectiveness<br>Safety<br>Person-centredness |
| EU Joint Action on Chronic Diseases and Healthy Ageing Across the Life Cycle (2019) [77] | Europe | QCR tool | Effectiveness<br>Safety<br>Person-centredness |
| Institute of Medicine (US) Committee on Quality of Health Care in America (1999) [78] | USA, but also used worldwide | None | Effectiveness<br>Safety |
| Peikes, et al. (2014) [79] | USA | evaluation | Timeliness, accessibility<br>Continuity |
| McDonald et al. (2014) [80] | USA | Care Coordination<br>Measurement Framework | Effectiveness<br>Safety<br>Person-centredness<br>Continuity |
| Thompson (undated) [81] | USA | Disease Management Program | Effectiveness<br>Person-centredness<br>Continuity |
| U.S Department of Health and Human Services<br>(2010) [82] | USA | None | Effectiveness<br>Person-centredness |
| Primary Health Care Advisory Group (2015) [83] | Australia | Health Care Home | Effectiveness<br>Safety<br>Person-centredness<br>Accessibility<br>Continuity |
| Paulus et al., eds (2012) [84] | Belgium | None | Effectiveness<br>Person-centredness<br>Continuity |
| Jackson et al. (2016) [85] | Canada | Continuity of Care | Continuity |
| Department of Health and Children (2008) [86] | Ireland | None | Effectiveness<br>Patient / person-centredness<br>Continuity |
| Quality Assurance Research and Policy<br>Development Group (2004) [87] | Philippines | Quality Standards | Effectiveness<br>Safety<br>Person-centredness<br>Accessibility, timeliness<br>Continuity |

More detailed information extracted from the scientific and grey literature can be found in the supplementary files, available from https://www.itg.be/en/research/research-themes/quality-of-care-for-chronic-conditions.

### Delphi survey

Forty nine of the 52 invited stakeholders (94%) consented and participated in the Delphi survey. Table 3 provides demographic and pertinent characteristics.

**Table 3.** Demographic characteristics of Delphi respondents (n=49)

|  |  |  |
| --- | --- | --- |
| Continent of origin | Africa | 10 |
|  | Asia | 12 |
|  | Europe | 11 |
|  | North America | 7 |
|  | Oceania | 2 |
|  | South America | 3 |
|  | Chose not to disclose | 3 |
|  | No answer | 1 |
| Socio-economic classification of country/ies of 'expertise' / having knowledge of | Low-income (LIC) | 10 |
|  | Middle-income (MIC) | 10 |
|  | High-income (HIC) | 3 |
|  | All | 7 |
|  | Both LIC and MIC | 13 |
|  | Both MIC and HIC | 3 |
|  | Not applicable | 3 |
| Stakeholder characteristics (multiple answers possible) | Clinician / health care provision | 15 |
|  | Health financing | 29 |
|  | Policy implementation | 19 |
|  | Policy formulation | 21 |
|  | Government adviser | 33 |
|  | Teacher or researcher in chronic conditions | 17 |
|  | Teacher or researcher in quality of care | 24 |
|  | Teacher or researcher in health financing | 20 |
|  | Informal caregiver of PwCC | 5 |
|  | PwCC | 4 |
|  | Civil society representative | 2 |
|  | Healthcare organization representative | 8 |
|  | Patient group representative | 4 |

### Specifying aims for good quality chronic care

For quality of care for chronic conditions, we noted that the six aims as proposed by the IOM: effectiveness; efficiency; safety; equitability; accessibility, timeliness, and affordability; and person-centredness also apply. Additionally, we identified a seventh aim, continuity of care. There was consensus among the Delphi participants on our propositions; they also provided reasons why each proposed aim should be included. One panellist recommended giving enough attention to integration to organise achieving the aims.

***<u>Effectiveness</u>*** is defined by the IOM [15] as the provision of services based on scientific knowledge to all who could benefit and refraining from providing services to those not likely to benefit (i.e., avoiding both overuse of inappropriate care and underuse of effective care). ‘Effectiveness’ was noted in 43 out of the 48 scientific articles[18–44,49–55,57–65] and in 24 of the 26 grey literature [3,4,12,14–15,67–78,80–84,86–87]. It is the most used quality aim, customarily measured with favourable (clinical) outcomes. Studies tend to utilise good clinical outcomes to demonstrate good quality of care or successful implementation of any model for chronic care. This is supported by various models of chronic care, where the “endpoints” are good clinical (and functional) outcomes.

This outlook seems supported by the Delphi respondents:

> *“we need…services/ interventions which are effective (yielding intended results)”*

and that there is

> *“no point in continuing if (care is) ineffective.”*

***<u>Efficiency</u>*** is a quality aim in 20 of the scientific articles [26,28,30,32,39,43–49,51–52,54–55,57,61,62,65] and in 9 of the grey literature[3,4,12,14–15,68–70,72]. This aim is about using appropriate (amounts of) resources and avoiding waste, including waste of equipment, supplies, ideas, and energy [12]. Efficiency is effectuated, for instance, by: reducing inappropriate use of emergency departments[28] and avoidable hospital admissions [32]; efficient use of time [57]; coordinating additional or specialised services only for PwCC who need these, etc. [12,85]. It also includes cost-efficient management that would lead to cost reductions without compromising beneficial effects [66], for instance the appropriate use of health technologies and information technology, coordinating additional or specialized services only for PwCC who need them, etc. [71].

A respondent highlighted cost-efficiency and related this to affordability:

> *“Efficiency combines effectiveness at a more appropriate cost; this can already solve many access issues, namely financial access issues.”*

***<u>Safety</u>*** is defined by the IOM as avoiding harm to PwCC from the care that is intended to help them [14,78]. The WHO [12] specifies this as ‘patient safety’. However, the grey literature from the Philippines [87] gives a broader goal *to provide patients, staff, and other individuals within the health facility a safe environment*. This quality aim appeared in only three of the scientific literature we reviewed [29,39,52] and in 10 of the grey literature [12,14,15,70,76–78,80,83,87].

A respondent indicated that safety can be complex and affected by the context.

> *“…given the context in which the implementation takes place, with low or middle (income country) organizational conditions lacking material and sometimes motivation from the workforce, safety can be a problem, especially in remote areas…”*

***Equity***, expressed as ensuring that all PwCCs can access good quality healthcare that is responsive to their needs regardless of personal characteristics [12] were identified in four of the scientific articles [23,29,40,43] and four other grey literature [3,14,15,70].

A respondent indicated that:

> *“Inequity has always been a major challenge, not only due to socioeconomic classes, but also due to clientelism practiced by politicians, including health ministers, whereby ’connected’ persons had better access to healthcare.”*

***<u>Accessibility, timeliness, and affordability</u>***, defined as reducing unwanted waits and harmful delays for both those who receive and those who give care, reducing access barriers and financial risk for patients, families and communities and promoting affordable care for the system, are presented in full or as one/two of the component-aims in 20 of the scientific papers we have reviewed [21,29,31,38–39,42–43,45–47,49–50,52,56–59,60–61,64], and implied in two more [27,40]. The full aim is presented in two grey literature [15,70] and as one/two of the component-aims in four more [14,79,83,87].

Respondents spoke about geographic and financial accessibility and timeliness, and linked accessibility to equity.

> *“The biggest challenge is access to care for chronic diseases, such as diabetes, for all patients wherever they are, given the geographical complexity (of specific countries).”*

> *“…there is a vast gap between the theoretical coverage of the social health insurance system and the actual coverage, which generates disparities in access to health care in general, particularly for people suffering from chronic conditions. This mainly affects people in rural areas and low-income communes in urban settings.”*

> *“Patients should (be able to) access quality chronic care when they need, without any barrier.”*

> *“…ensure that services of health providers are generally accessible to all and equitably distributed.”*

***<u>Person-centredness</u>*** is defined by WHO [70] as the approach to care that consciously adopts the perspectives of individuals, carers, families and communities as participants in, and beneficiaries of, trusted health systems organized around the comprehensive needs of people rather than individual diseases, and respects social preferences. This is a central dimension in 36 of the scientific papers [18,20,29,31–39,41–44,48–53,58,62–65] and ranges from outright mentions of patient-/person-centredness to implicit indications that relate to collaborating with and engaging PwCC in their care and self-management. It is also included in 19 more grey literature [3–4,12,14–15,68,71,73–77,80–84,86–87] we reviewed.

In the Delphi, respondents connected person-centredness to access and talked about ‘empowerment’.

> *Within a context of rapidly escalating co- and multi-morbidities, it is critical (to) reflect (on) mechanisms which create access to care for the "person" and not the "disease or conditions".*

> *I would put a premium on empowering patients to be able to do self-care, particularly for interventions that have been proven to be effective.*

We identified a 7^th^ aim, “continuity of care”, which is particularly important for people who require regular, consistent healthcare services for a long period of time. Continuity is among the recommendations by the Institute of Medicine in 2001 [14] (*Care based on continuous healing relationships*). The WHO’s ICCC Framework [73] extensively discusses continuity across time and care settings**. *<u>Continuity</u>*** is the focus of one of our grey literature (Jackson et al.[85]), where it is defined as the *degree to which a series of discrete health events is experienced as coherent and connected, and consistent with the patient’s healthcare needs and personal context.* It includes the capacity to monitor and respond to change, support self-management goals, and link to community resources [37] as well as follow-up and tracing of lost-to-follow-up [23,57]. Three sub-dimensions are given by Jackson et al.[85], which we utilised as further basis for inductive and deductive analysis:

> [i] Relational/relationship continuity, defined as *a trusting relationship with one or more HCP who help bridge healthcare episodes over time.*

> [ii] Informational continuity, where health information as well as other relevant information about the PwCC (their values, preferences, contexts) are shared (i.e., shared medical records).

> [iii] Management continuity, where patient-related information regarding their case management is communicated to different relevant HCP.

Continuity is mentioned in 14 other grey literature [3,4,12,68,71,73–74,79–81,83–84,86–87]. It is also explicitly included as a quality dimension in five [23,29,37,40,49] of the scientific literature we retained and implied in 17 more [19–21,28,31,33,37,39,41–42,51–52,56,57,59,61,63].

Respondents placed value on continuity as a quality aim, especially for chronic care.

> *“…continuity of care is probably the major issue regarding chronic diseases…”*

> *“A lack of continuity of care is currently one of the main reasons patients who access the system fall through the cracks.”*

> *“Continuum of care is vital for NCDs…”*

Based on our definition, ‘***Integration***’ does not qualify as an aim. We note that some of our grey literature refers to it as a measurable characteristic of quality of care (i.e., considered as an aim)[12] or as a necessary characteristic of (people-centred) health care [84]. One grey literature focuses on integration, where it is described as an action or a process [4]. Thirteen scientific literature [24,39–41,43,44,46,49,51,61–63,65] we reviewed mention it as instrumental to improving quality of care for chronic conditions.

One of the Delphi respondents indicated:

> *“a key thing is the principle (the ‘what’), and that is the principle of integrated chronic care. Right now, we have reactive, short-term care… changing that mindset is priority.”*

Another stated that

> *“for improved care on chronic conditions we need to achieve… delivery within integrated care pathways spanning across (the) care sector and being organised around the patient…”*

## DISCUSSION

Because they last for a long time and, more often, throughout the lifetime of the person, chronic conditions raise particular issues in healthcare. Another consideration is the natural history of chronic conditions and the ‘journey’ of a PwCC through time, traversing the natural history. To respond to these, healthcare services would encompass risk- and disease-prevention, clinical management of the condition and any complications and/or multi-morbidity, rehabilitation and/or community re-integration, palliative/end-of-life care, and considering the psychosocial aspects of the PwCC. Additionally, clinical/biomedical control of chronic conditions is not static. PwCCs experience episodes of good and poor control of the condition at different moments throughout their lifetime. Control of the condition can be affected by many factors such as continued exposure to risks and determinants, other health problems, e.g., infections, psychological issues (anxiety, depression, etc.), co-morbidity, suboptimal clinical management by either the healthcare system or by the PwCC/informal caregivers themselves, as well as social factors (e.g., lack of social support). These redound to specific healthcare needs and corresponding services and supports the view to reflect on how the quality of such healthcare would be defined.

We have formulated a definition and determined quality aims paying attention to the specificities of chronic conditions and its care. We define quality of chronic care *as the degree to which healthcare services for individuals and populations with chronic conditions – including provision of education and support to adapt and self-manage in the face of social, physical, and emotional challenges – which are consistent with current professional knowledge and increase the likelihood of desired health outcomes and biopsychosocial well-being.* This complements our chronic care quality aims, wherein achievement of each is influenced by and would contribute to realization of our definition.

Regarding the aims, we noted from the literature review and the Delphi survey that most of the IOM care quality aims take on new, additional meanings specific for chronic care.

Further to ‘effectiveness’ as a chronic care quality aim is the consideration that people develop chronic condition(s) due to exposure to various risk factors and social determinants of health [18,20]. These would also still affect the person even if they already have a chronic condition, and increase the propensity of having poor clinical control, emergence of complications and/or development of other morbidities [30]. Thus, effective health services to prevent and control risks, e.g. healthy lifestyle promotion, smoking cessation counselling, etc., should be provided to the general population and the PwCCs (in addition to their effective case management). This was also pointed out by our Delphi survey respondents. Logically, failure to provide effective services to prevent and control risks would increase the number of PwCCs and could give rise to multi-morbidity. Compounding this with failure to deliver effective chronic care would further increase the burden of chronic conditions. However, an understanding of the responsibilities of the health system needs to be clarified. Addressing the various risks and determinants themselves, e.g., air pollution control, increasing access to healthy food, regulation of sales of unhealthy products, food (re)formulations, tobacco and alcohol taxations, etc. would need actions beyond the scope of the health system, even if the health system may initiate multi-sectoral policies and actions [89]. Furthermore, effectiveness should not be limited to chronic care provision alone. PwCC are often immunocompromised, making them susceptible to various infections. Responsive health systems should thus have the capacity and capability to effectively provide care for both chronic conditions and acute infections and support the psychosocial needs of PwCC.

With the ballooning burden of chronic conditions and higher demands for chronic care, efficiency becomes a dire necessity across all settings. Whilst efficiency can be looked at individual level (best use of own resources), it has more relevance as an aim at collective level. From the societal perspective, efficiency ensures that what is not wasted on one person is available for the other who truly needs it. Efficiency, especially of chronic care, is one of the main aims of many recent health system reforms, which are, however, being implemented mainly in high-income countries.

We echo one of our Delphi respondents that safety is indeed complex and largely depends on the context. Moving from concentrating only on patient safety, we favour the more extensive safety aim presented in the PhilHealth Benchbook [87], which includes measures for both patients and (healthcare) staff and providing a safe environment for healthcare delivery. In addition, effects of chronic care delivery on the environment should also be considered; for instance, cytostatic drugs excreted by cancer patients finding their way to wastewater and affecting biodiversity of freshwater organisms. Concrete examples of broader safety are: having appropriate facility design, e.g. to accommodate people with disabilities; prevention of adverse care incidents, e.g. drug-induced hypoglycaemia, drug-drug interactions in cases of polypharmacy (especially among PwCC with comorbidities/multi-morbidity); provision of equipment and devices needed to deliver safe care, e.g. personal protective equipment; proper waste handling, including proper disposal of sharps used by both the staff and PwCC, appropriate wastewater treatment, etc. While safety was the least studied quality aim among the papers we reviewed, its importance in chronic care and its wider application including effects on global environmental changes cannot and should not be discounted.

Equity is a cross-cutting consideration in our societies. It can be assessed over any metric of interest and consists in a normative judgement on the distribution within a group of persons; it is usually not an aim at individual level, but at collective level. For instance, there can be equity considerations on how households financially contribute to the general funding of the health system, with different views on what would be fair. Health systems performance and UHC literature has put ***<u>equity</u>*** as a core consideration [70]. Akin to equity in healthcare in general and as defined by the IOM, we propose to refer to equity as the aim capturing distributional considerations related to quality of chronic care. The first concern should be that care does not vary in quality (effectiveness, safety, timeliness, person-centredness, etc.) because of personal characteristics such as gender [64], ethnicity, socioeconomic status, disabilities, etc [29]. Such fairness redounds in health promotion and all levels of prevention of chronic conditions, especially in LMICs where exposure to risks and the prevalence of most chronic conditions have been documented to be higher among lower socio-economic groups, and where more inequities have also been noted (e.g. between sexes, ethnicity).

As regards individual PwCC, we deduce, and view, person-centredness to be more holistic and more all-encompassing than what is defined by both IOM and WHO. To deliver person-centred chronic care, what should be strived for should go beyond the “*patient is a person*” concept. The recognition and acceptance should be framed as: the “*person is sometimes a patient*” and the "*patient is always a person*”. The biomedical, psychological and social aspects of PwCC need to be considered [10,12,50,52]. They should be “activated”/stimulated to become experts of their condition and should be guided to accept and recognise that their chronic condition is only one of the facets of their whole life. Emphasis is given on their *human agency* and their capacity to make their own decisions, set goals, take actions, and sustain efforts. Therefore, beyond empowerment/enablement for self-management, there should be engagement of the PwCC for collaborative care and involvement in applicable committees to improve chronic care quality [25,42,71]. Such engagement extends to their family, their informal caregivers and the community [23,39,49,63]. A goal would be that PwCCs will be able to “juggle” the different facets of their life effectively to maximize their physical/biological, psychological and social well-being, and are supported accordingly.

**Accessibility, affordability, and timeliness** are mainly instrumental to other aims. However, as argued below, they also have intrinsic value. Following Levesque et al.’s [88] proposition, we further dissect this aim as applied to chronic care:

(a) Geographic accessibility – to help enable PwCC to consult and follow-up regularly without having to travel a great distance and/or encountering much difficulties in traveling and/or losing much time for travel and/or incurring unreasonable transportation expenses [29,61]. It is a source of reassurance: for PwCCs to know that in case of need, there is a solution nearby. Less uncertainty reduces stress; it also expands choices for daily life.

(b) Financial accessibility (*affordability)* of services, medications and supplies – as too-high and/or recurring financial costs charged to the PwCCs compromise their welfare and may force PwCC to stop their maintenance medications, (e.g. cannot buy high-priced insulin) or cause *iatrogenic poverty* (e.g. need to go into debt to finance chemotherapy) [29,47,49,52]. It also frees resources for other needs.

(c) Temporal accessibility – with considerate opening hours, and reasonable waiting times (for example for consultations) and turn-around-times (e.g. of laboratory results)[21,38,39,47] as long waiting times for laboratory test results or to receive appropriate care can create distress and anxiety, if not the worsening of the condition.

(d) Availability of chronic care services, including diagnostics and when, i.e. periodic or consistent, and availability of medications [27,31,42–43,46,49–50,56,60,64] – considering that making services and diagnostics available either every day or at regular specific schedules would be optimal for a PwCC who will need to utilise these services repeatedly; as well as

(e) health care worker-related factors, e.g. cultural congruence, approachability [36,46,49,63–64] – as these highly contribute to acceptability, prompting better access, and building trust.

The effects of achieving the different sub-dimensions of the above aim are many-fold: from improving chronic care delivery and utilisation of healthcare services, contributing to improved adherence of PwCC, improving (clinical) outcomes, etc. Achieving this aim would directly contribute to the achievement of good quality chronic care, as we defined.

The intrinsic value of our seventh aim, continuity, is how it acknowledges that time and its continuity (past, present and future) matters for the PwCC. It addresses chronicity, which can be discerned from the three subdimensions proposed by Jackson et al [85].

[i] Relational/relationship continuity, as establishing rapport and trust between the HCP and the PwCC would be crucial to build a lasting relationship, help ensure regular follow-up, and more likely promote (long-term) adherence [29,39,51–52].

[ii] Informational continuity, to ensure timely availability of the health information of PwCCs across different HCP as they navigate different healthcare disciplines, different levels of care, through time, including any changes of HCP (e.g. when PwCC relocate)[19–21,28–29,31,33,37,39,41–42,49,56,63]; and

[iii] Management continuity, where patient-related information regarding their case management is communicated to relevant HCP and shared with the PwCC themselves and/or their informal caregivers. This way, care is delivered across different health sectors and by multiple HCP as well as by the PwCC themselves and/or their informal caregivers in a coherent, logical and timely fashion [19,21,28,31,33,37,39,41–42,49,56,61,63].

Informational and management continuity go hand-in-hand, where relevant information about the PwCC (informational continuity) as well as care management plans (management continuity) would be co-developed and shared with relevant care providers (including informal caregivers) and the PwCC and would follow the PwCC in their journey through healthcare.

A thorny issue is the status of ‘integration’, for which multiple definitions have been given [4]. Certainly, poor and insufficient integration of service delivery is a major issue in many countries given the current state of their health systems. This observation, shared across settings including in high-income countries, probably motivated the decision to elevate integration as an aim in another quality of care framework [12]. However, we propose to distinguish the conceptual undertaking of identifying the aims of quality of care from policy agendas inspired by the current situation. From a conceptual perspective, our assessment is that integration does not meet the intrinsic value criterion of an aim. It, however, plays a special role in care quality as it is instrumental to other aims. The process of integration or the action of integrating healthcare organizations and/or its people can help achieve quality care aims, more particularly effectiveness, efficiency, person-centeredness, and continuity. Furthermore, integration is not a directional metric (for which any progress is valuable). Conceptually, integration is desirable, but only to the extent that it positively serves the aims care quality. Empirically, we acknowledge that it should be a top priority in all health systems; a way to organise actions to improve quality of (chronic) care.

## LIMITATIONS

Available literature on quality of (chronic) care mostly documents experiences in high-income countries, this was also the case in our scoping review. We mitigated this limitation by purposively selecting Delphi participants with expertise and experiences on chronic care in LMICs.

We also noted that specific aims got more attention over others (for instance, effectiveness was the focus of almost all of the scientific literature we reviewed while less than ten were on equity, safety). Although this may be an unintended effect of our literature search (especially in the choice of data bases), we noted that effectiveness is usually the *de facto* aim that is studied because of the value placed on desired clinical outcomes as the endpoint. However, this does not warrant inattention to the other aims. The inclusion of grey literature, most of which tackled all aims, made sure that we have a broad understanding even of the least studied aims in the scientific literature.

We have adopted the view that ‘aims’ should be intrinsically valuable. It is important to understand that this does not mean that non-included aims are irrelevant.

Although we introduced the PwCC journey and the relevance of the various stages in a continuum of care relevant to the identified aims, we are aware that we did not have room to expound more on this concept. We will discuss this in more detail in a separate paper presenting our conceptual quality of chronic care framework.

## CONCLUSIONS

With this paper, we have moved from a generic understanding of quality of care to one tailored to chronic conditions. Beyond aims, we have also determined the scope of attention, one which values a comprehensive offer of healthcare services, addresses risks and social determinants of health, ensures biopsychosocial well-being of PwCC, and gives importance to quality of care characteristics relevant to the PwCC and their families, to the community, and to the health system.

Our scoping review shows that some aims have received more attention than others. However, limited attention should not be interpreted as an acceptable reason for neglect. Our Delphi survey respondents underlined the value of each of the seven chronic care quality aims that we identified, especially as applied to low-income settings.

## IMPLICATIONS FOR POLICY & FURTHER RESEARCH

Our team used these aims to create a chronic care quality framework, and then, via the Delphi survey, mobilized the international panel of experts to apply the said framework for possible purchasing arrangements to improve quality of chronic care in low- and middle-income countries. The chronic care quality framework and the Delphi survey results on purchasing arrangements will be presented separately. These are all components of the larger program of work implemented by WHO, which focuses on purchasing arrangements as an instrument to improve health services for chronic conditions. It is expected that member nations will take inspiration from this program of work, in their efforts to improve care for chronic conditions. Actors active in chronic care may also be inspired by our specifications, in designing good quality chronic care services or working on improvement strategies thereto.

The output we presented in this paper is conceptual. Operationalization for systematic improvements in quality of chronic care can be a next step, among others, to demonstrate the usefulness, or not, of each of these specified chronic care quality aims, in specific settings.

Our paper may also inspire other calibrations and validation of the definition and aims of quality of care for other health problems. We hypothesise that they could be valuable preparatory steps among those committed to improve quality of care for specific health conditions. Having a tailored understanding of quality of care will only make quality improvement interventions better.

## Data Availability

All data produced in the present study are available upon reasonable request to the authors

## Acknowledgments

We thank John de Maesschalck, Institute of Tropical Medicine, Antwerp, Belgium, Rafael Nalupta and Shreyashi Paik (student interns) for their assistance with the screening of scientific literature in the scoping review. The online Delphi survey (administration) was facilitated by Lynette Dominguez, independent consultant and supported, as for the digital platform, by Martin Erpicum, Mesylab, Liege, Belgium.

We are grateful to the participants of our Delphi panel for their invaluable contributions.

## Author Contributions

GK conceptualized the full work, based on a terms of reference published by WHO, with contributions from WVDP, MGAA, and DK. The scoping review protocol was prepared by GK, with contributions from WVDP, MGAA, DK, MR and BM. The scoping review was conducted by GK, WVDP, MGAA, and DK. The Delphi survey protocol was prepared by GK and BM, with contributions from WVDP, MGAA, DK, and MR. The Delphi survey questionnaire was prepared by BM and GK, with contributions from MR. Analysis of the Delphi survey was conducted by BM, GK and MR. This manuscript was drafted by GK, with contributions from WVDP, MGAA, DK, MR and BM. All authors read and approved the final version.

